# Separating Signal from Noise in Wastewater Data: An Algorithm to Identify Community-Level COVID-19 Surges

**DOI:** 10.1101/2022.09.19.22280095

**Authors:** Aparna Keshaviah, Ian Huff, Xindi C. Hu, Virginia Guidry, Ariel Christensen, Steven Berkowitz, Stacie Reckling, Rachel T. Noble, Thomas Clerkin, Denene Blackwood, Sandra McLellan, Adélaïde Roguet, Isabel Mussa

**Author notes:** Corresponding author Aparna Keshaviah.

## Abstract

Wastewater monitoring has shown promise in providing an early warning for new COVID-19 outbreaks, but to date, no approach has been validated to reliably distinguish signal from noise in wastewater data and thereby alert officials to when the data show a need for heightened public health response. We analyzed 62 weeks of data from 19 sites participating in the North Carolina Wastewater Monitoring Network to characterize wastewater metrics before and around the Delta and Omicron surges. We found that, on average, wastewater data identified new outbreaks four to five days before case data (reported based on the earlier of the symptom start date or test collection date). At most sites, correlations between wastewater and case data were similar regardless of how wastewater concentrations were normalized, and correlations were slightly stronger with county-level cases than sewershed-level cases, suggesting that officials may not need to geospatially align case data with sewershed boundaries to gain insights into disease transmission. Wastewater trend lines showed clear differences in the Delta versus Omicron surge trajectories, but no single wastewater metric (detectability, percent change, or flow-population normalized viral concentrations) adequately indicated when these surges started. After iteratively examining different combinations of these three metrics, we developed a simple algorithm that identifies unprecedented signals in the wastewater to help clarify changes in communities’ COVID-19 burden. Our novel algorithm accurately identified the start of both the Delta and Omicron surges in 84% of sites, potentially providing public health officials with an automated way to flag community-level COVID-19 surges.

## Introduction

Wastewater monitoring has emerged as a promising tool to measure SARS-CoV-2 and other population health markers in communities (1, 2). The method yields objective, anonymous data on the nearly 80% of U.S. households connected to a sewer system (3). For COVID-19, this approach also overcomes biases and shortcomings in clinical testing data stemming from several factors, including reliance on healthcare seeking behavior, unequal access to diagnostic tests (4–6), delays in clinical test result reporting (7), lack of routine testing of asymptomatic individuals, who account for 40% to 45% of COVID-19 infections (8), and imprecise case attribution due to reporting by permanent residence rather than residence at the time of testing (9). Further, as a population-level data source, wastewater provides a more cost-effective way to detect the presence of new viral variants circulating in a community compared to individual testing and sequencing (10). Going forward, wastewater data may become a valuable *independent* signal of COVID-19 infections as the shift to at-home testing further reduces public health officials’ visibility into community infections.

A recent survey conducted to assess the role of wastewater data in pandemic management revealed that one of the features of greatest interest to public health officials is the potential of wastewater monitoring to raise an early warning for SARS-CoV-2 entry or spread in a community (11). However, the “lead time” advantage of wastewater data remains difficult to pinpoint, in part because it may vary across different phases of a disease outbreak. Early in the pandemic, studies comparing trends in wastewater viral concentrations with trends in clinical case counts reported lead times ranging from two days (12, 13) to two to three weeks (14, 15). More recent research has shown that using the specimen collection date rather than reporting date for case data shortens the wastewater lead time (16), and that when clinical tests are widely available and widely used, wastewater and clinical data are well-aligned, with no lead-time advantage of one over the other (6). The variability in reported lead times stems from a combination of biological early detection in wastewater (“latent” factors) and reporting lags in the clinical tests (“pragmatic” factors) (17, 18). Further, site to site differences in lead times can arise from differences in laboratory analytical methods, sewer infrastructure (including sewage travel time), and testing behaviors in the population.

Importantly, the early warnings that wastewater data can provide come not only from its potential to be a leading indicator, but also from the ability to monitor a much broader swath of the population than is typically tested clinically. However, population-level data are only useful to the extent that public health officials can interpret the data and use it to inform pandemic response (11). While states have implemented different approaches to alerting officials to changes in COVID-19 risk based on the wastewater data (19), there is a dearth of research validating these approaches, and at present, no algorithms or metrics have been identified to reliably distinguish signal from noise in wastewater data.

To help fill these knowledge gaps, we analyzed longitudinal data from the statewide North Carolina Wastewater Monitoring Network (NCWMN) to demonstrate how states can better use wastewater data to get early warnings of new outbreaks and variant-based surges. We first compared wastewater viral concentrations to reported county- and sewershed-level case counts to assess the lead-time advantage of wastewater data. In doing so, we examined how results varied across sites with different wastewater treatment plant (WWTP) service population sizes, and whether different approaches to normalizing wastewater viral concentrations (to adjust for sample-to-sample differences in wastewater flow rate, the size of the contributing population, and lab assay sensitivity) meaningfully altered our findings. We then characterized spatial and temporal patterns in wastewater metrics leading up to surges resulting from the Delta and Omicron variants. Lastly, we developed and validated a multi-metric algorithm to distinguish signal from noise in the wastewater data and identify the start of the Delta and Omicron surges in each site.

## Results

We analyzed data from 1,783 wastewater samples collected from January 3, 2021 to March 15, 2022 from 19 WWTPs in 16 counties, along with clinical case data from these counties. The WWTPs included in our analysis span the state from west to east (Figure S1) and vary in service population size, covering 3,500 and 550,000 people, or 5% to 60% of the county’s resident population (Table 1). The frequency and timeframe of wastewater sample collection varied by site.

**Table 1.** Characteristics of North Carolina wastewater treatment plants analyzed.

| Site name | County | County population | WWTP service population (percent of county) |  | Sampling timeframe | Number of samples |
| --- | --- | --- | --- | --- | --- | --- |
| Laurinburg | Scotland | 34,823 | 15,527 | (45%) | 6/16/21–3/14/22 | 72 |
| Tuckaseegee | Jackson | 43,938 | 13,296 | (30%) | 1/3/21–3/14/22 | 94 |
| Marion | McDowell | 45,756 | 8,459 | (18%) | 6/16/21–3/14/22 | 73 |
| Beaufort | Carteret | 69,473 | 3,500 | (5%) | 1/4/21–3/15/22 | 123 |
| Roanoke Rapids | Halifax | 69,493 | 14,320 | (21%) | 6/18/21–3/15/22 | 73 |
| City of Wilson | Wilson | 81,801 | 49,384 | (60%) | 6/18/21–3/14/22 | 71 |
| Chapel Hill – Carrboro | Orange | 148,476 | 78,141 | (53%) | 1/5/21–3/15/22 | 117 |
| Greenville | Pitt | 180,742 | 89,616 | (50%) | 1/4/21–3/15/22 | 121 |
| Wilmington City (North) | New Hanover | 234,473 | 58,361 | (25%) | 1/4/21–3/15/22 | 114 |
| New Hanover County (North) | New Hanover | 234,473 | 67,743 | (29%) | 1/21/21–3/15/22 | 114 |
| South Durham | Durham | 321,488 | 108,105 | (34%) | 1/5/21–3/15/22 | 118 |
| Fayetteville – Rockfish | Cumberland | 335,509 | 151,589 | (45%) | 6/18/21–3/8/22 | 73 |
| MSD of Buncombe County | Buncombe | 378,608 | 173,000 | (46%) | 6/18/21–3/15/22 | 69 |
| Winston Salem – Salem | Forsyth | 382,295 | 178,000 | (47%) | 6/18/21–3/15/22 | 70 |
| Greensboro – North Buffalo | Guilford | 537,174 | 135,821 | (25%) | 6/17/21–3/15/22 | 75 |
| Charlotte 1 | Mecklenburg | 1,110,356 | 68,685 | (6%) | 1/4/21–3/14/22 | 111 |
| Charlotte 2 | Mecklenburg | 1,110,356 | 182,501 | (16%) | 1/3/21–3/14/22 | 113 |
| Charlotte 3 | Mecklenburg | 1,110,356 | 120,000 | (11%) | 5/31/21–3/14/22 | 74 |
| Raleigh | Wake | 1,111,761 | 550,000 | (49%) | 1/5/21–3/15/22 | 108 |

Note: Analyses were limited to sites with sustained monitoring around the Delta and Omicron surges. Sites are ordered by ascending county population size.
MSD = Metropolitan Sewerage District; WWTP = wastewater treatment plant.

### Early warning potential: wastewater versus clinical data

To examine the lead-time advantage of wastewater data, we compared trends in wastewater viral concentrations (normalized by flow rate and service population size) with trends in clinical case counts (recorded based on the symptom start date or test specimen collection date, whichever was earlier). We examined correlations with both county-level and sewershed-level case counts. Although neither comparison is perfect, we prioritized the former, recognizing that wastewater data may capture the infections of not just those who reside in sewershed, but also those who are unsewered but travel to sewered areas for work or recreation.

We found that rises in wastewater viral concentrations leading up to the Delta and Omicron surges generally coincided with the timing of rises in county-level case counts (Figure 1a,1b). However, around the Omicron surge, wastewater viral concentrations peaked more quickly than case count data in most sites (Figure 1b).

**Figure 1.**
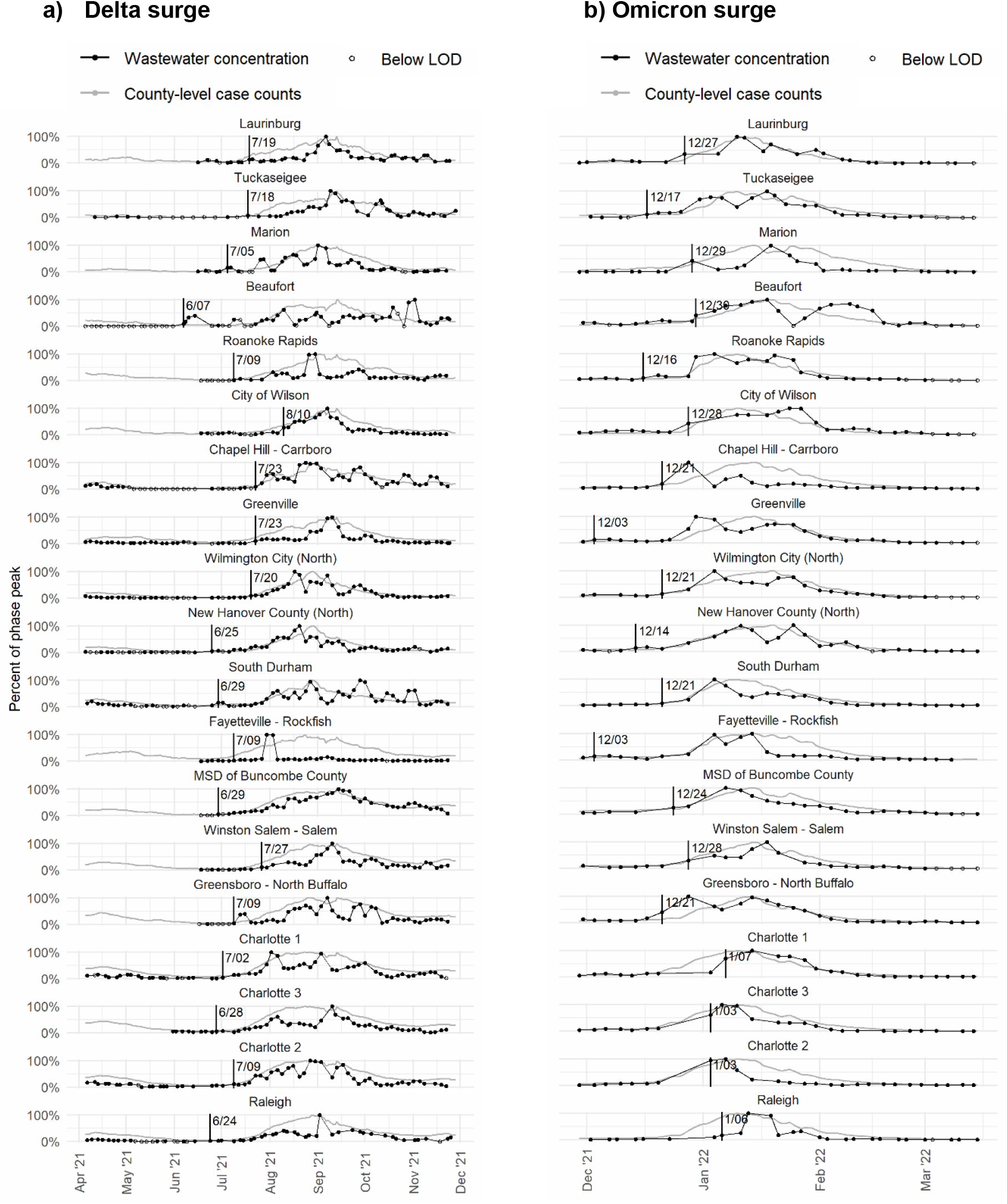
Trends in wastewater viral concentrations and COVID-19 case counts. Note: Wastewater viral concentrations (based on the arithmetic means of the N1 and N2 concentrations) were normalized by flow rate and service population size. County-level case counts are based on 7-day rolling averages. Vertical lines mark the start of each surge (based on visual inspection of wastewater trend lines). Sites are ordered by ascending county population size. LOD = limit of detection; MSD = Metropolitan Sewerage District.

In most sites, correlations between flow-population normalized wastewater viral concentrations and case counts (both county-level and sewershed-level) were strengthened when we lagged the case data—that is, when we compared wastewater concentrations on day *x* with case counts on day *x+lag* (Figure 2, Table 2, Table S1)—suggesting that wastewater tends to be a leading indicator. In 12 of 19 sites, the lead time that maximized the correlation with county-level case counts was 4 to 7 days but ranged from −1 day (indicating that the wastewater data lagged the case data) to 12 days across all sites (Table 2).

**Table 2.**
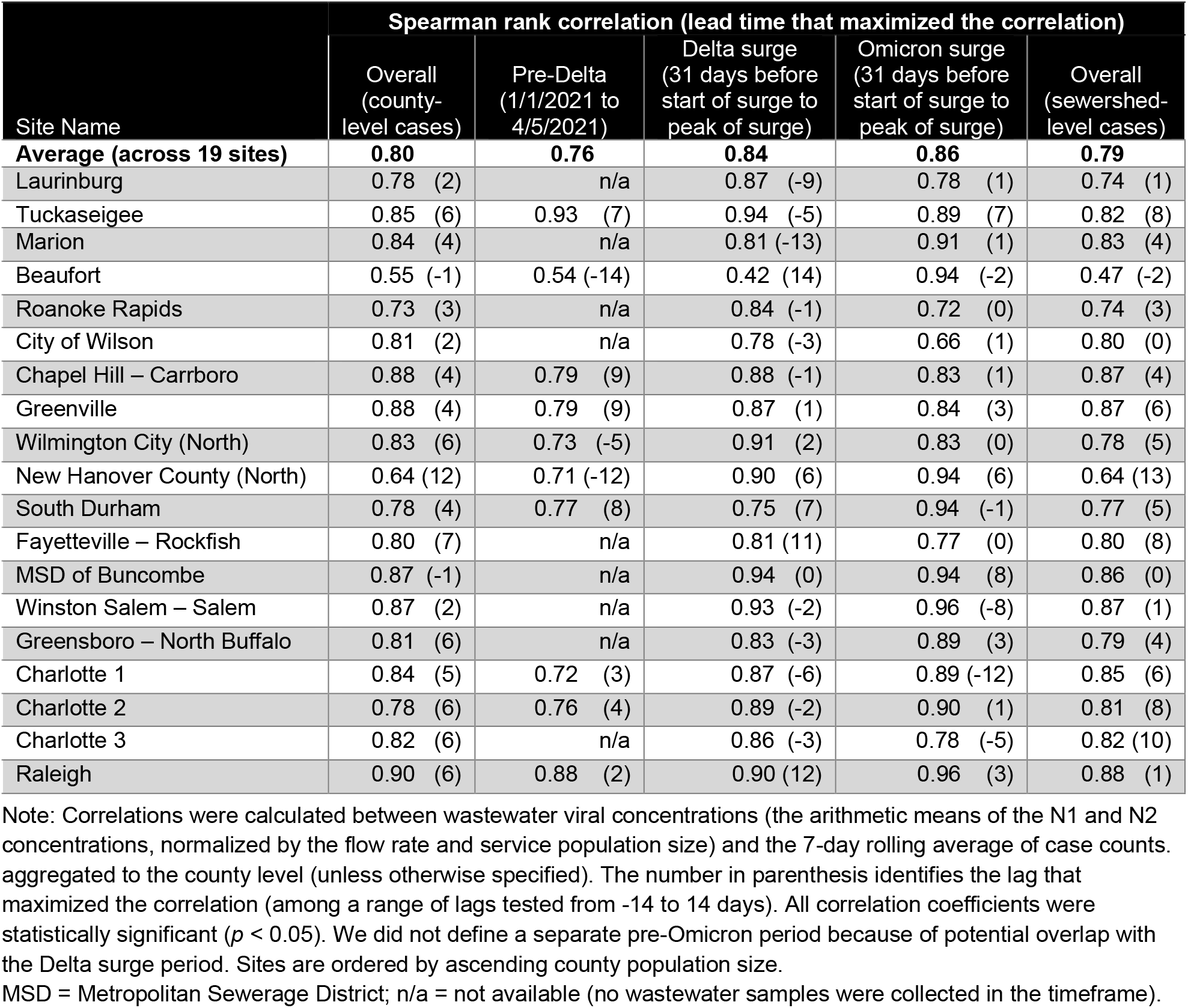
Correlations between wastewater viral concentrations and lagged case counts.

| Site Name | Spearman rank correlation (lead time that maximized the correlation) |  |  |  |  |
| --- | --- | --- | --- | --- | --- |
|  | Overall (county-level cases) | Pre-Delta (1/1/2021 to 4/5/2021) | Delta surge (31 days before start of surge to peak of surge) | Omicron surge (31 days before start of surge to peak of surge) | Overall (sewershed-level cases) |
| **Average (across 19 sites)** | **0.80** | **0.76** | **0.84** | **0.86** | **0.79** |
| Laurinburg | 0.78 (2) | n/a | 0.87 (-9) | 0.78 (1) | 0.74 (1) |
| Tuckaseegee | 0.85 (6) | 0.93 (7) | 0.94 (-5) | 0.89 (7) | 0.82 (8) |
| Marion | 0.84 (4) | n/a | 0.81 (-13) | 0.91 (1) | 0.83 (4) |
| Beaufort | 0.55 (-1) | 0.54 (-14) | 0.42 (14) | 0.94 (-2) | 0.47 (-2) |
| Roanoke Rapids | 0.73 (3) | n/a | 0.84 (-1) | 0.72 (0) | 0.74 (3) |
| City of Wilson | 0.81 (2) | n/a | 0.78 (-3) | 0.66 (1) | 0.80 (0) |
| Chapel Hill – Carrboro | 0.88 (4) | 0.79 (9) | 0.88 (-1) | 0.83 (1) | 0.87 (4) |
| Greenville | 0.88 (4) | 0.79 (9) | 0.87 (1) | 0.84 (3) | 0.87 (6) |
| Wilmington City (North) | 0.83 (6) | 0.73 (-5) | 0.91 (2) | 0.83 (0) | 0.78 (5) |
| New Hanover County (North) | 0.64 (12) | 0.71 (-12) | 0.90 (6) | 0.94 (6) | 0.64 (13) |
| South Durham | 0.78 (4) | 0.77 (8) | 0.75 (7) | 0.94 (-1) | 0.77 (5) |
| Fayetteville – Rockfish | 0.80 (7) | n/a | 0.81 (11) | 0.77 (0) | 0.80 (8) |
| MSD of Buncombe | 0.87 (-1) | n/a | 0.94 (0) | 0.94 (8) | 0.86 (0) |
| Winston Salem – Salem | 0.87 (2) | n/a | 0.93 (-2) | 0.96 (-8) | 0.87 (1) |
| Greensboro – North Buffalo | 0.81 (6) | n/a | 0.83 (-3) | 0.89 (3) | 0.79 (4) |
| Charlotte 1 | 0.84 (5) | 0.72 (3) | 0.87 (-6) | 0.89 (-12) | 0.85 (6) |
| Charlotte 2 | 0.78 (6) | 0.76 (4) | 0.89 (-2) | 0.90 (1) | 0.81 (8) |
| Charlotte 3 | 0.82 (6) | n/a | 0.86 (-3) | 0.78 (-5) | 0.82 (10) |
| Raleigh | 0.90 (6) | 0.88 (2) | 0.90 (12) | 0.96 (3) | 0.88 (1) |

Note: Correlations were calculated between wastewater viral concentrations (the arithmetic means of the N1 and N2 concentrations, normalized by the flow rate and service population size) and the 7-day rolling average of case counts aggregated to the county level (unless otherwise specified). The number in parenthesis identifies the lag that maximized the correlation (among a range of lags tested from -14 to 14 days). All correlation coefficients were statistically significant ( $p < 0.05$ ). We did not define a separate pre-Omicron period because of potential overlap with the Delta surge period. Sites are ordered by ascending county population size.
MSD = Metropolitan Sewerage District; n/a = not available (no wastewater samples were collected in the timeframe).

**Figure 2.**
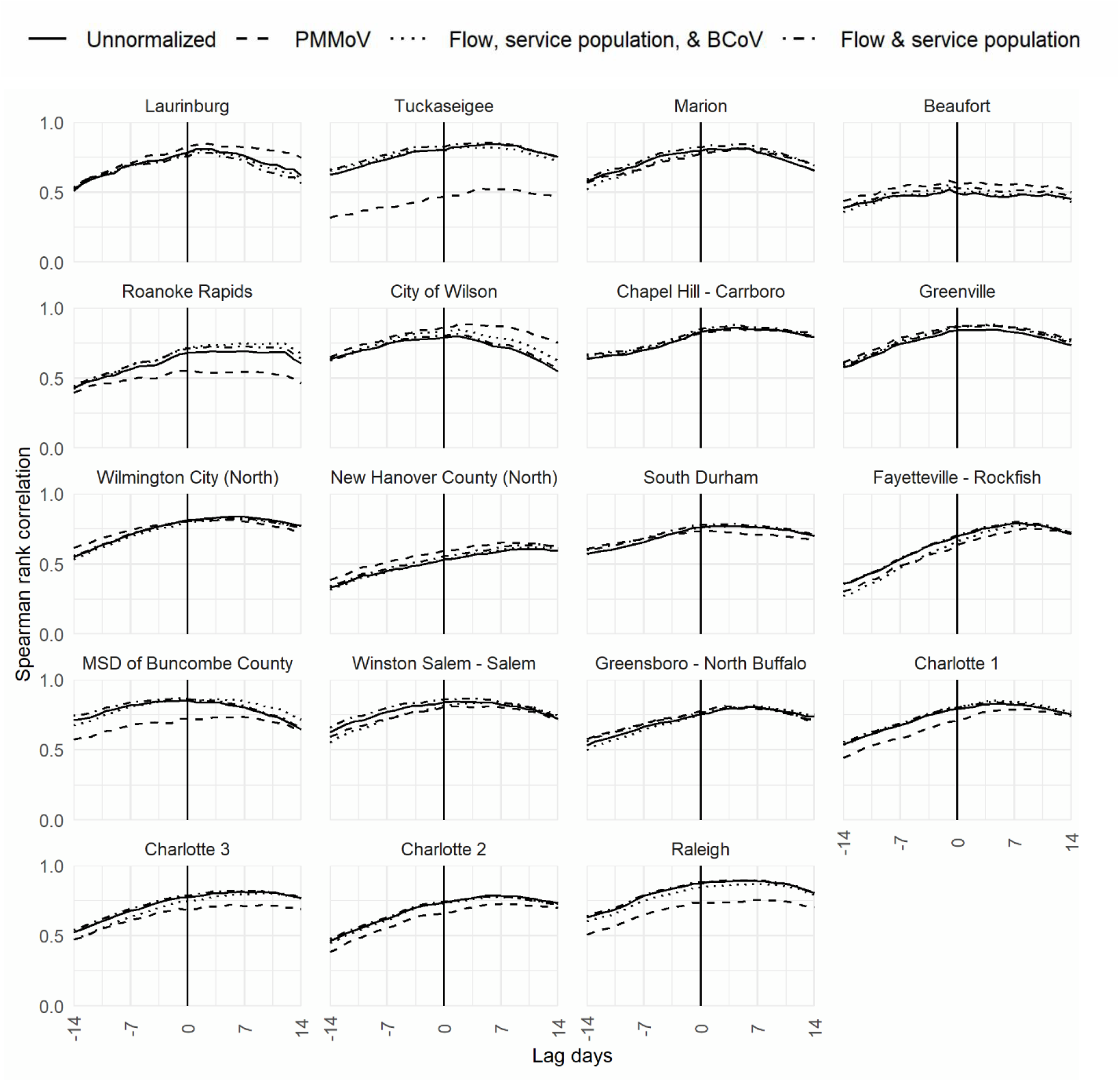
Correlations between COVID-19 case counts and wastewater viral concentrations, by wastewater normalization method. Note: Correlations were calculated between wastewater viral concentrations (the arithmetic means of the N1 and N2 concentrations), normalized using different approaches, and the 7-day rolling average of county-level case counts. Sites are ordered by ascending county population size. BCoV = Bovine coronavirus; PMMoV = Pepper mild mottle virus.

In analyses to examine the generalizability of our findings, we found the following:

- The lead-time advantage of wastewater data was somewhat shortened when compared to sewershed-level case counts than county-level case counts, and wastewater viral concentrations were slightly less correlated with sewershed-level case counts than county-level case counts (averaging 0.79 versus 0.80 and ranging from 0.47 to 0.87 versus 0.55 to 0.90, respectively), though differences were not meaningful (Table 2).
- Restricting the data to periods around the Delta and Omicron surges (that is, excluding periods of low case counts prior to the Delta surge) generally increased the strength of the correlation between wastewater data and case counts, though differences in the strength of correlation varied by site and pandemic phase (Table 2). The lead-time advantage of wastewater data also fluctuated greatly across sites within a period, and across periods within a site. For example, in Wilmington City (North), wastewater concentrations lagged case counts during the pre-Delta period (with a −5-day lead time maximizing the correlation), lead case counts during the Delta surge (with a 2-day lead time maximizing the correlation) and were aligned with case counts during the Omicron surge (with neither source leading the other). Around the Delta surge, the correlation averaged 0.84 across sites (range: 0.42 to 0.94), was slightly higher around the Omicron surge, averaging 0.86 across sites (range: 0.66 to 0.96).
- The average correlation between wastewater viral concentrations and sewershed-level case counts was higher among sites with WWTPs that served larger populations (*r* = 0.84 among plants serving 144,000 to 550,000 people) than smaller populations (*r* = 0.75 among plants serving 3,500 to 32,500 people).
- Normalizing wastewater viral concentrations did not substantially impact the lead-time advantage of the wastewater data, and in most sites, different normalization approaches yielded a similar level of correlation with case counts across the range of lead and lag times examined (Figure 2). However, in some sites (including Tuckaseigee, Raleigh, Roanoke Rapids, and MSD of Buncombe County), the correlation between county-level case counts and wastewater viral concentrations was diminished when the wastewater concentrations were normalized by pepper mild mottle virus (PMMoV) concentrations rather than by flow rate and WWTP service population size; flow rate, WWTP service population size, and bovine coronavirus (BCoV) recovery rate; or unnormalized.

### Patterns in wastewater metrics leading up to the Delta and Omicron surges

Historical trends in wastewater viral concentrations captured the differing trajectories of the Delta and Omicron surges. On average, the time from the start of the Delta surge (based on visual inspection of site-specific low-population normalized wastewater trend lines) to its peak was 55 days but ranged from 21 days (in Fayetteville – Rockfish) to 144 days (in Beaufort, the smallest site included in statewide surveillance, which had some of the noisiest wastewater data over time). During the Omicron surge, wastewater viral concentrations rose more quickly and drastically; the average time from the start of the surge to its peak was only 18 days and ranged from 0 days (in Charlotte 1, where the peak was represented by a single-sample spike followed by a steady decline thereafter) to 42 days (in New Hanover County (North)). Generally, at the start of the Delta and Omicron surges, the percent increase in wastewater viral concentrations was much higher in smaller sites (based on county population or sewershed population size) than in larger sites. Below, we describe wastewater metrics around these surges in more detail.

#### Delta surge

The rise and fall of wastewater viral concentrations just before and at the start of the Delta surge varied greatly across the 19 sites (Figure 1). In the month before the surge began, 58% of samples had detectable concentrations, and wastewater viral concentrations were rising in some sites and falling in others, yielding an average percent change of 5% (Table 3). In the 10-day window before the start of the Delta surge, 69% of samples had detectable wastewater viral concentrations, and concentrations increased by an average of only 3%. At the start of the Delta surge at each site, wastewater metrics changed drastically. All samples had detectable concentrations, and viral concentrations increased by an average of 706%. In absolute terms, flow-population normalized viral concentrations increased from an average of 1.3 million and 1.2 million viral copies per person per day in the 31-day and 10-day period before the start of the surge, respectively, to an average of 7.1 million viral copies per person per day across sites at the start of the surge.

**Table 3.** SARS-CoV-2 wastewater concentration metrics leading up to surges.

| Site Name | Percent change in normalized wastewater viral concentrations |  |  |  |  |  |
| --- | --- | --- | --- | --- | --- | --- |
|  | Delta surge |  |  | Omicron surge |  |  |
|  | Average over prior month | Average over prior 10 days | Value at start of surge | Average over prior month | Average over prior 10 days | Value at start of surge |
| **Average (across 19 sites)** | **5%** | **3%** | **706%** | **77%** | **43%** | **768%** |
| Laurinburg | 103% | 17% | 1665% | 51% | -8% | 1108% |
| Tuckaseegee | undef | -76% | undef | 8% | 16% | undef |
| Marion | 75% | 163% | 1198% | 70% | 1% | 2,530% |
| Beaufort | undef | undef | undef | 27% | 27% | 129% |
| Roanoke Rapids | undef | undef | undef | 152% | -42% | 3,034% |
| City of Wilson | 16% | -29% | 2075% | 26% | 11% | 3,465% |
| Chapel Hill – Carrboro | -66% | -13% | undef | 123% | 150% | 311% |
| Greenville | -29% | 81% | 354% | 61% | 148% | 110% |
| Wilmington City (North) | 62% | 29% | 267% | 44% | 11% | 226% |
| New Hanover County (North) | -67% | n/a | 669% | 32% | -85% | 150% |
| South Durham | -77% | undef | 513% | 23% | 14% | 80% |
| Fayetteville – Rockfish | 252% | -9% | 800% | 81% | 34% | 108% |
| MSD of Buncombe | undef | undef | undef | 522% | -31% | 207% |
| Winston Salem – Salem | -46% | -57% | 596% | 27% | 18% | 188% |
| Greensboro – North Buffalo | -61% | undef | undef | 61% | 99% | 131% |
| Charlotte 1 | -57% | -29% | 501% | 30% | 208% | 575% |
| Charlotte 2 | 38% | -55% | 215% | 61% | n/a | 597% |
| Charlotte 3 | -13% | 26% | 303% | 53% | n/a | 701% |
| Raleigh | -75% | n/a | 19% | 91% | 159% | 181% |
| **Normalized wastewater viral concentrations**<br>**(copies per person per day, in millions)** |  |  |  |  |  |  |
| Site Name | Delta surge |  |  | Omicron surge |  |  |
|  | Average over prior month | Average over prior 10 days | Value at start of surge | Average over prior month | Average over prior 10 days | Value at start of surge |
| **Average (across 19 sites)** | **1.25** | **1.20** | **7.06** | **8.44** | **9.68** | **68.94** |
| Laurinburg | 3.52 | 2.32 | 16.94 | 10.35 | 8.41 | 101.60 |
| Tuckaseegee | 0.94 | 0.30 | 4.09 | 4.39 | 2.36 | 50.61 |
| Marion | 1.64 | 1.04 | 19.60 | 4.07 | 5.65 | 150.83 |
| Beaufort | 0.37 | 0.45 | 4.45 | 8.83 | 15.82 | 45.63 |
| Roanoke Rapids | 0.36 | 0.37 | 3.43 | 11.97 | 8.18 | 32.68 |
| City of Wilson | 2.30 | 3.89 | 31.15 | 8.25 | 6.00 | 57.03 |
| Chapel Hill – Carrboro | 0.30 | 0.64 | 2.23 | 4.47 | 9.41 | 42.78 |
| Greenville | 1.50 | 1.45 | 8.48 | 3.99 | 4.65 | 20.05 |
| Wilmington City (North) | 0.77 | 1.18 | 5.78 | 6.07 | 3.88 | 16.29 |
| New Hanover County (North) | 0.23 | n/a | 2.30 | 4.86 | 3.40 | 16.57 |
| South Durham | 0.51 | 0.84 | 5.17 | 3.23 | 4.64 | 10.08 |
| Fayetteville – Rockfish | 3.01 | 1.25 | 8.78 | 4.80 | 11.17 | 25.66 |
| MSD of Buncombe | 0.16 | 0.22 | 1.68 | 5.85 | 8.05 | 24.74 |
| Winston Salem – Salem | 0.58 | 0.43 | 3.02 | 7.37 | 9.97 | 28.71 |
| Greensboro – North Buffalo | 0.26 | 0.17 | 2.06 | 8.14 | 18.50 | 59.65 |
| Charlotte 1 | 0.77 | 0.38 | 2.27 | 10.70 | 20.35 | 137.34 |
| Charlotte 2 | 2.38 | 1.59 | 5.02 | 14.48 | n/a | 183.10 |
| Charlotte 3 | 3.48 | 3.13 | 6.90 | 22.82 | n/a | 174.09 |
| Raleigh | 0.33 | 0.73 | 0.87 | 13.57 | 47.10 | 132.33 |

Note: Wastewater viral concentrations are based on mean of N1 and N2 concentrations, normalized by the flow rate and service population size. The percent change was calculated as the sample-to-sample change in the normalized wastewater viral concentrations. Values are reported from 31 days before the identified start of the surge (based on visual inspection of normalized wastewater trend lines) to the start of the surge at that site.
MSD = Metropolitan Sewerage District; n/a = not available (no samples were collected in the timeframe shown);
undef = undefined (prior samples had concentrations below the limit of detection that were not precisely quantified).

While wastewater metrics averaged across all 19 sites clearly changed at the start of the Delta surge (which was based on site-specific start dates, rather than a given calendar date), we found that no single metric could reliably identify the start of the surge at each site. Instead, considering multiple metrics in combination and in the context of historical trends was helpful to identify the start of the surge. We also found that the percent change can be a misleading metric when a site has low wastewater concentrations that hover around the limit of detection (LOD), since a 3- to 6-fold increase may still result in a relatively low concentration. Indeed, focusing on percent change alone resulted in false positives in some sites, but false negatives in others. For example, in Laurinburg, wastewater concentrations increased by a larger percent in the month prior to the start of the surge (103%) than in the 10 days leading up to the surge (17%). Visual inspection of the trend line shows that the higher percent increase in the month prior was driven by a single sample that was followed by a sustained period of lower concentrations. In Raleigh, the percent increase in the wastewater viral concentration at the start of the Delta surge was not very high (at 19%), but in absolute terms, the viral concentration at the start of the surge hit a level that had not been seen for more than 7 weeks. Further, the percent increase occurred after a period in which wastewater concentrations were either declining or below the LOD.

#### Omicron surge

Patterns in wastewater metrics around start of the Omicron surge varied from patterns around the Delta surge, likely because Delta infections had not fully waned by the time the Omicron variant first surfaced in the state. In the month before the Omicron surge began, 98% of samples had detectable concentrations (compared to 58% prior to the Delta surge), and wastewater viral concentrations increased by an average of 77% (Table 3). In the 10-day period before the start of the Omicron surge, 94% of samples had detectable concentrations, and wastewater viral concentrations increased by an average of only 43%. The declining wastewater viral concentrations that many sites saw during this period (compared to the month prior) likely reflects a waning of Delta infections before the start of the Omicron surge. At the start of the Omicron surge, the wastewater metrics again changed drastically. All samples had detectable concentrations and wastewater viral concentrations increased by an average of 768%. On absolute terms, flow-population normalized viral concentrations increased from an average of 8.4 million and 9.7 million viral copies per person per day in the 31-day and 10-day period before the Omicron surge, respectively, to an average concentration of 68.9 million viral copies per person per day across sites at the start of the surge.

### An algorithm to distinguish signal from noise in wastewater viral concentrations

Given our findings that no single metric summarizing wastewater viral concentrations (detectability, percent change, or normalized viral concentrations) reliably identified the start of the Delta and Omicron surges, we iteratively tested different sets of logic, based on multiple wastewater metrics, in combination to identify the start of the Delta and Omicron surges more accurately.

Figure 3 describes the logical criteria that best distinguished signal from noise in the wastewater data to identify a surge start date that aligned with our gold standard start date (based on visual inspection of trends in site-specific flow-population normalized wastewater viral concentrations).

**Figure 3.**
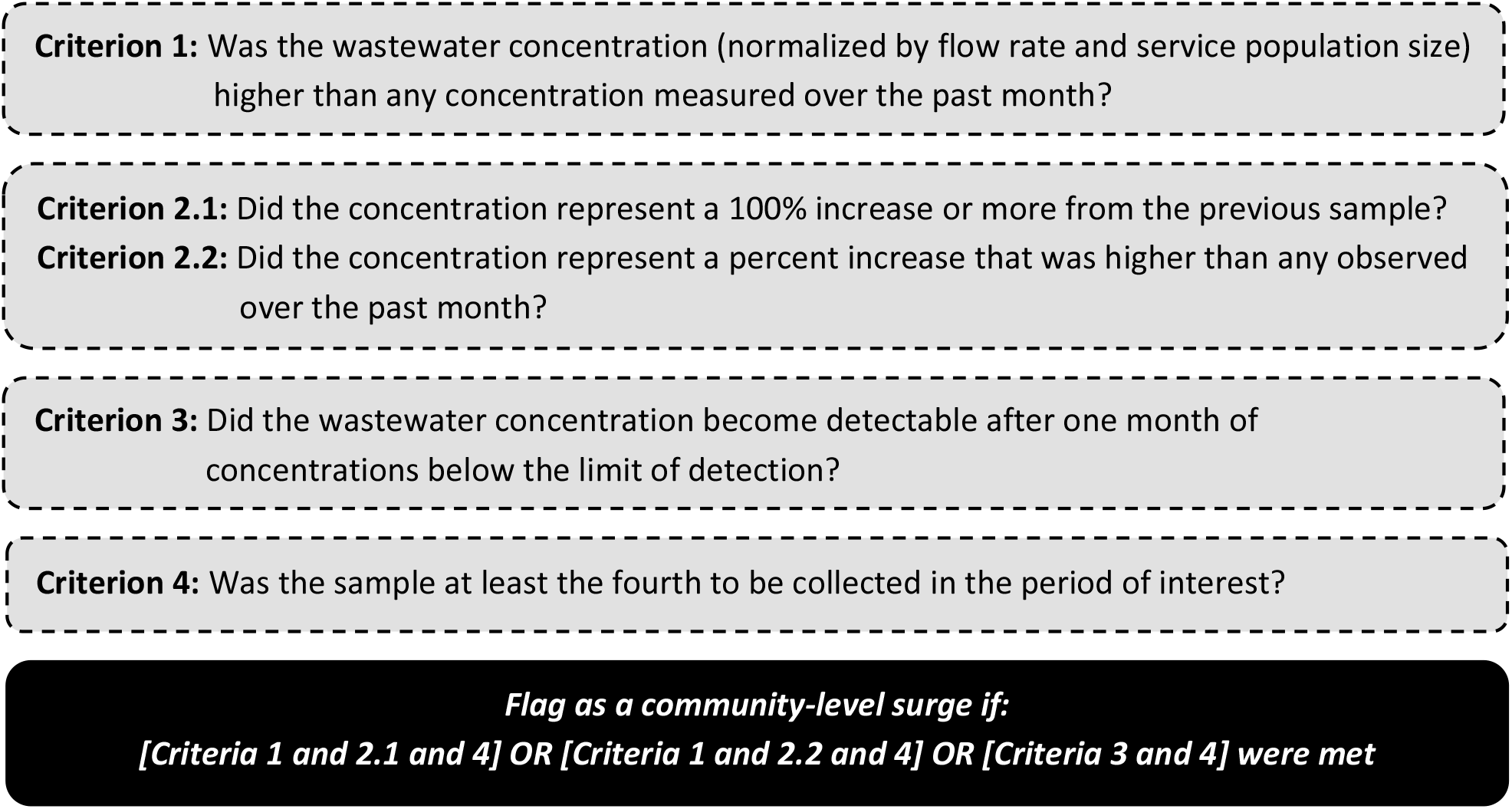
An algorithm to flag community-level surges using wastewater data.

After applying to logical criteria to identify all wastewater samples for which an alert was flagged, we selected the earliest flagged sample within each period of interest (that is, after the Delta variant was first identified in the state, and after the Omicron variant was first identified in the state, based on retrospective sequencing of clinical samples) to determine the algorithmically identified Delta and Omicron surge start dates. We then compared the algorithmically identified start dates with the start dates we had identified based on visual inspection of trends to assess performance (Table 4).

**Table 4.** Surge start dates based on visual inspection of trends versus a novel algorithm.

| Site Name | Delta surge start date (algorithm) | Delta surge start date (visual inspection) | Omicron surge start date (algorithm) | Omicron surge start date (visual inspection) |
| --- | --- | --- | --- | --- |
| Laurinburg | 07/19/2021 | 07/19/2021 | 12/27/2021 | 12/27/2021 |
| Tuckaseegee | 07/05/2021† | 07/18/2021† | 12/31/2021† | 12/17/2021† |
| Marion | 07/05/2021 | 07/05/2021 | 12/29/2021 | 12/29/2021 |
| Beaufort | 06/07/2021 | 06/07/2021 | 12/29/2021 | 12/30/2021 |
| Roanoke Rapids | 07/09/2021 | 07/09/2021 | 12/16/2021 | 12/16/2021 |
| City of Wilson | 07/06/2021† | 08/10/2021† | 12/28/2021 | 12/28/2021 |
| Chapel Hill – Carrboro | 06/25/2021† | 07/23/2021† | 12/21/2021 | 12/21/2021 |
| Greenville | 06/07/2021† | 07/23/2021† | 12/28/2021† | 12/03/2021† |
| Wilmington City (North) | 06/29/2021† | 07/20/2021† | 12/21/2021 | 12/21/2021 |
| New Hanover County (North) | 06/25/2021 | 06/25/2021 | 12/14/2021 | 12/14/2021 |
| South Durham | 06/29/2021 | 06/29/2021 | 12/21/2021 | 12/21/2021 |
| Fayetteville – Rockfish | 07/09/2021 | 07/09/2021 | 12/21/2021† | 12/03/2021† |
| MSD of Buncombe | 06/29/2021 | 06/29/2021 | 12/24/2021 | 12/24/2021 |
| Winston Salem – Salem | 07/27/2021 | 07/27/2021 | 12/28/2021 | 12/28/2021 |
| Greensboro – North Buffalo | 07/13/2021† | 07/09/2021† | 12/17/2021† | 12/21/2021† |
| Charlotte 1 | 07/02/2021 | 07/02/2021 | 12/17/2021† | 01/07/2022† |
| Charlotte 2 | 07/09/2021 | 07/09/2021 | 01/03/2022 | 01/03/2022 |
| Charlotte 3 | 06/18/2021† | 06/28/2021† | 12/17/2021† | 01/03/2022† |
| Raleigh | 06/24/2021 | 06/24/2021 | 12/16/2021† | 01/06/2022† |

† Denotes differences in the surge start date when based on the algorithm versus visual inspection of wastewater trend lines (normalized by flow rate and service population size).

Summarizing the alignment between the two approaches, and thus the performance of the algorithm (Table 5), we found the following:

**Table 5.** Performance of algorithm versus manual identification of variant-related surges.

| Period | Comparable<br>(accurate start) | Superior<br>(early warning) | Unclear<br>(false alarm or<br>micro-surge) | Inferior<br>(missed surge) |
| --- | --- | --- | --- | --- |
| Delta (n=19) | 63% | 21% | 11% | 5% |
| Omicron (n=19) | 63% | 21% | 0% | 16% |
| **Both surges (n=38)** | **63%** | **21%** | **5%** | **11%** |

Note: Rows may not sum to 100% due to rounding error. Predictive value of the algorithm (grey-shaded columns) was calculated by combining sites where algorithm had identical or superior performance to manual identification.

- The algorithmically identified surge start date **coincided with** the visually identified start date (±1 day) in 12 of 19 sites during the Delta surge and in 12 of 19 sites during the Omicron surge—i.e., in 24 of the 38 classification periods.
- The algorithm performed **better than** visual inspection, picking up smaller perturbations in wastewater viral concentrations that were unprecedented but not visually obvious, in 4 sites during the Delta surge and 4 sites during the Omicron surge. The early warning ranged from 10 to 28 days during the Delta surge and 4 to 21 days during the Omicron surge. Notably, in Charlotte 3, the algorithm identified an Omicron surge start date that was just before a two-week gap in wastewater monitoring around the winter holidays—in other words, it would have provided officials with an early warning of the surge, based on an unprecedented value that may not have been obvious at the time based on visual inspection of the trend line.
- The algorithm had **unclear performance** in 2 sites during the Delta period (by 35 and 46 days), but no sites during the Omicron period. In both sites (Greenville and City of Wilson), the algorithm flagged a single sample with an elevated viral concentration that was followed by several samples with lower concentrations. Here, it’s unclear if the flagged “blip” reflects a false alarm (i.e., underlying variability in wastewater measurements that was not associated with a change in cases) or a micro-surge (i.e., an actual increase in cases that simply did not result in sustained community transmission).
- The algorithm **missed** the start of the surge for 1 site during the Delta period (by 4 days) and 3 sites during the Omicron period (by 14 to 25 days). The missed start during the Delta period occurred because the sample collected at the true start of the surge had a detectable viral concentration that was preceded by 2.5 weeks of concentrations below the LOD (not the 1-month period required in criterion 3). In two of the three sites where the start of the Omicron surge was missed, the true start of the surge occurred between 0-4 days after the Omicron variant was first known to have entered the state (and so the sample did not meet the requirements of criterion 4), while in the third site, the percent change metric was undefined at the true start of the surge because the prior sample had a concentration below the LOD (and so the sample did not trigger criteria 2.1 or 2.2).

Overall, our algorithm accurately identified or provided an early warning for the start of the Delta and Omicron surges in 32 of 38 classification periods, yielding a predictive value of 84%. The predictive value increased to 89% if we presume that in the two sites with unclear performance, the algorithm flagged micro-surges (rather than false alarms). When we based calculations on unnormalized wastewater viral concentrations instead of flow-population normalized concentrations, algorithm performance was slightly weakened: the algorithm missed the start of the surge in 6 of the 38 classification periods (versus 4 when based on flow-population normalized concentrations).

The criteria shown in Figure 3 are listed in general order of importance. Criterion 3 was useful in smaller sites (like Beaufort) where many samples were below the limit of detection. Criterion 4 was implemented to establish a baseline level of noise and thereby reduce false alarms, though in some sites, it led to a missed start date (note: the 4-sample cut-off corresponded to an observation period of at least 8 days in North Carolina). Finally, for the one-month period referenced, we found that use of a 33- to 34-day window optimized the algorithm’s performance, though performance was still good when we applied a 28- to 32-day window.

## Discussion

The findings from our analysis of data from North Carolina’s statewide wastewater monitoring program validate the use of wastewater monitoring to assess patterns in community transmission of COVID-19. Temporal trends in wastewater viral concentrations aligned well with trends in reported case counts, and wastewater data reflected the differing trajectories of the Delta and Omicron surges. Wastewater data appeared to be a leading indicator in most sites, identifying community outbreaks an average of four to five days before (county- and sewershed-level) clinical case data overall, and one to two days before the major variant-based surges. Notably, we observed an uptick in the wastewater data fully one week before the first Omicron case was announced in the state (20), and in different parts of the state (in Greenville and Fayetteville – Rockfish, rather than Charlotte). We also found that no single wastewater metric adequately identified the Delta or Omicron surge. However, by looking at the proportion of samples with detectable levels, the percent change in smoothed wastewater viral concentrations, and absolute wastewater viral concentrations together, we were able to sharpen focus around changing community risk profiles.

The results of our analyses provide several practical insights for how public health officials can use wastewater data. First and foremost, we were able to develop a multi-metric algorithm that reliably distinguished signal from noise to accurately identify the start of both the Delta and Omicron surges in 84% of sites. At present, NCWMN weekly wastewater reports include two metrics: percentiles (based on the magnitude of wastewater viral concentrations) and percent change (over a 15-day period). For those unfamiliar with wastewater data, such metrics can be difficult to interpret, as they do not provide a basis for understanding when public health officials should take action to curb COVID-19 transmission. Indeed, as we note above, percent change was often misleading, as several sites saw a 100% increase or more in wastewater concentrations (the highest classification, coded in red on the statewide dashboard) when viral concentrations were quite low and hovering around the LOD. The algorithm we developed helps fill this gap, and because it is based on logic that can be easily implemented with spreadsheet software, it can be readily incorporated into existing dashboards to alert officials to when wastewater data signals a need for heightened public health response. Further, based on ongoing analyses, our algorithm shows promise to generalize outside of North Carolina and accurately flag variant-based surges in other states.

Second, we found that correlations were similar when we compared wastewater viral concentrations to county-level versus sewershed-level case counts, which aligns with findings from Duvallet et al (21). Our results suggest that officials can make valid comparisons in near real time with county-level case data and may not need to geospatially align wastewater data with WWTP sewersheds to gain insights into disease transmission patterns. This finding is important because many WWTPs lack shapefiles delineating the geospatial boundaries of their service populations, and the analytic processing needed to restrict case count data to the sewershed can be cumbersome and infeasible for public health agencies that lack staff with expertise in geospatial methods.

Third, given that the lead-time advantage of wastewater data fluctuated greatly across sites within a period (possibly due to differences in WWTP size or clinical test result turnaround time) and across periods within a site (possibly due to changes in the number of people getting tested, and in transmissibility and infection characteristics as circulating variants changed), wastewater data analysis and reporting may need to be tailored to site features (such as WWTP service population size), the overall infection rate, and the characteristics of the circulating pathogen or variant. Developing processes for adaptive wastewater metric development and refinement can help public health officials stay on top of new threats as they arise.

Our findings also extend the literature in several ways. Whereas previous reports from statewide wastewater programs have focused more on urban communities with large WWTPs (16, 21), North Carolina’s statewide program includes rural communities with small WWTPs (the smallest of which serves 3,500 people). The underrepresentation of small WWTPs may explain why Duvallet et al found no relationship between the strength of correlation between PMMoV normalized wastewater and clinical case data and WWTP service population size (21). Our finding that normalizing wastewater levels did not substantially alter correlations between wastewater and case data aligned with some previous studies, but not others. In two previous analyses that included data from several sites, findings were similar to our study (16, 21). But in past analyses that largely focused on data from a single site, correlations between wastewater and case data were improved after normalizing wastewater concentrations (22, 23).

There are a few limitations and assumptions that may have impacted our findings. First, because NCDHHS was not conducting wastewater-based variant monitoring at the time of this analysis, we could not confirm that observed rises in wastewater viral concentrations were due to the Delta or Omicron variants. However, when we dug further into wastewater data from the one site where we had variant monitoring in place—Jackson County—we saw that the date when wastewater variant sequencing results first detected the Delta variant, on July 5, coincided with our algorithmically-identified Delta surge start date (per Table 4). A second consideration is that the correlation results we present might be sensitive to our use of a 7-day rolling average to summarize clinical case counts. We chose this approach to smoothing to eliminate cyclic patterns in the case data, whereby no cases were recorded on weekends, and instead those cases were reflected in each Monday’s count. In sensitivity analyses, we found that the strength of correlation between wastewater viral concentrations and raw daily case counts (with no smoothing) was virtually unchanged, but that the lead time that maximized the correlation was attenuated to 2 to 3 days, on average, when no smoothing was applied. Finally, because wastewater monitoring takes place in only a minority of the 100 counties across the state, with sampling occurring twice weekly, on average (based on standards for participation in the Centers for Disease Control and Prevention’s National Wastewater Surveillance System), wastewater data are not yet granular enough to assess patterns in geographic spread, particularly with threats that transmit rapidly, like the Omicron variant. More research is needed to determine optimal sampling schemes for sentinel public health warnings.

The algorithm we introduce here is part of a broader effort to translate wastewater lab results into meaningful information that can directly inform public health management. By considering patterns across multiple wastewater metrics in combination, we were able to distinguish signal from noise in wastewater data and automate the identification of the start of two very different COVID-19 surges due to different viral variants. Our algorithm gives public health officials a timely way to assess the need for action, and the logical criteria we lay forth provide a framework for states to finetune the alert algorithm to their data.

## Materials and Methods

### Wastewater data

In January 2021, the North Carolina Department of Health and Human Services (NCDHHS) launched the NCWMN to pilot routine wastewater monitoring for SARS-CoV-2, as one of eight initial states funded by the Centers for Disease Control and Prevention’s National Wastewater Surveillance System. The NCWMN, which expanded the work of the NC Wastewater Pathogen Research Network (24), was funded by the state legislature at University of North Carolina (UNC) system institutions in 2020, and has grown to include 25 sites, with plans for continued expansion in Fall 2022. Nineteen sites were actively part of the network when this analysis was conducted. Wastewater samples collected through NCWMN, based on twice-weekly 24-hour time- or flow-weighted composite sampling at centralized WWTPs, were sent to a UNC laboratory for digital PCR analysis. Detailed UNC wastewater testing methods have been previously published (25).

Predating the pilot and prior to NCDHHS’ expansion to include sites in the underserved rural mountainous part of the state, Mathematica had launched wastewater monitoring in Western North Carolina, with funding from Dogwood Health Trust, which was provided through the Jackson County Department of Public Health. Wastewater samples were collected by the Tuckaseigee Water & Sewer Authority (TWSA) and tested by the University of Wisconsin-Milwaukee’s School of Freshwater Sciences (UWM). A detailed description of UWM’s wastewater sample preparation and testing procedures can be found in previous publications (16, 26). After a four-week feasibility study in summer 2020 (not included in the current analysis), TWSA began collecting weekly samples (n = 116) beginning in mid-December 2020 from the largest of three WWTPs in the county using time-weighted (hourly) composite sampling. In August 2021, NCDHHS provided supplemental funding to increase sampling in Jackson County to occur twice weekly, and the site was included in the NCWMN.

From November 2021 onwards, NCDHHS supported all wastewater monitoring in Jackson County, and wastewater sample analysis was transitioned from UWM to UNC from January 13, 2022 – February 28, 2022. The procedures used for wastewater sample processing and analysis were largely aligned across the UNC and UWM labs, with two main exceptions (beyond differences in the reagents used by the labs): (1) UNC pasteurized samples prior to analysis, while UWM did not, and (2) UNC used a two-step process to quantify the SARS-CoV-2 virus, while UWM used a one-step PCR process (which, based on UWMN’s optimization processes, yielded increased assay sensitivity, but did not affect the estimated viral concentrations). To assess the impact of these differences, NCDHHS conducted an inter-lab validation study prior to the transition. Over the 8-week validation period, a moderate level of agreement was observed between the two labs. The Spearman rank correlation coefficient comparing the average of the N1 and N2 concentrations between the two labs was 0.76, and the ratio of the log-transformed average of the N1 and N2 wastewater viral concentrations ranged from 0.91 to 1.25. Since March 2022, UNC has been analyzing all samples collected through the NCWMN.

### Reported case-based surveillance data

For comparison with wastewater viral concentrations, we downloaded reported case counts from the publicly available NC COVID dashboard (https://covid19.ncdhhs.gov/dashboard/data-behind-dashboards), which records cases based on the test specimen collection date or indicated date of symptom start, whichever was earlier. We downloaded county-level daily new case counts, as well as sewershed-level case rates—that is, the number of daily new cases per 10,000 people among those who reside within the boundaries of the sewersheds (WWTP service regions) that NCDHHS monitors. Prior to calculating sewershed-level case rates, NCDHHS set sewershed-level case counts between 1 and 4 to a value of 2, to meet NCDHHS Division of Public Health data suppression guidelines. To estimate sewershed-level case counts for each WWTP, we multiplied the sewershed-level case rates by the size of the population served by that WWTP.

### Statistical methods

The wastewater viral concentrations we analyzed represent the arithmetic mean of N1 and N2 viral concentrations. Prior to analysis, we cleaned indicator variables for whether N1 or N2 concentrations were below the limit of detection (LOD) using information on the actual N1 and N2 concentrations and the N1 and N2 LOD thresholds reported by the labs. We then derived a composite indicator for whether *both* the N1 and N2 concentrations were below the LOD for a given wastewater sample (versus whether at least one target had a concentration above the LOD). We excluded from our analysis five wastewater samples that were flagged as having quality control issues. We normalized wastewater viral concentrations by flow rate and service population size (27) to create an interpretable metric in the units of genome copies per person per day (note: for one sample with a missing flow rate, we carried forward the flow rate from the previous sample).

To assess the lead-time advantage of wastewater data, we calculated the correlation between flow-population normalized wastewater viral concentrations and case counts after lagging and leading the case counts by up to 14 days in either direction (for example comparing a wastewater measurement on a given day with clinical case counts measured *x* days before and after that wastewater sample was collected). Specifically, we calculated a Spearman rank correlation coefficient between the two data sources across the sampling period for each site. For these analyses, we applied a 7-day rolling average to new daily case counts to avoid cyclic gaps in the data arising from the fact that case counts are not reported on weekends. We also used single-value imputation to (1) set wastewater measurements below the LOD (which were reported as 0 by the UNC lab) to a value equal to the LOD divided by the square root of 2, and (2) set sewershed-level case counts that were missing to a count of 0. For WWTPs that serve multiple counties, we summed county-level case counts across the relevant counties covered by the WWTP for comparison with the site-level wastewater viral concentrations.

To assess the generalizability of our findings, we examined how the correlation between wastewater viral concentrations and clinical case counts varied in several ways. We first compared correlations with both county-level and sewershed-level case counts. Next, because long periods with low infection rates could unduly influence the measured relationship between the wastewater and clinical case data, we repeated the correlation analyses after restricting the data to periods around each major surge, defined as 31 days prior to the start of each surge (defined by visual inspection of the wastewater trend line, as described below) through the peak of that surge (based on when the maximum wastewater concentration or 7-day rolling average of clinical case counts occurred, whichever was later). We also assessed how correlations differed when wastewater data were generated from large and small WWTPs, defined based on quartiles of the WWTP’s service population size. Lastly, we examined how results changed after applying different normalization approaches to the wastewater data, including normalizing by (1) flow rate and service population size, (2) flow rate, service population size, and BCoV recovery rate, and (3) PMMoV, a human biomarker that serves a proxy for how many people contributed to the sewer system (28).

For analyses characterizing wastewater metrics around the Delta and Omicron surges, we reported the absolute magnitude of flow-population normalized viral concentrations, sample-to-sample percent changes in those concentrations (calculated only for samples with detectable concentrations), and the proportion of samples with a concentration above the LOD. We summarized wastewater metrics on the start date of each surge, during a 10-day window prior to the start of each surge, and during a one-month (31-day) window prior to the start of each surge. We identified the start of the Delta and Omicron surges by conducting a visual assessment of the wastewater trend lines for each site. When plotting the trend lines, we narrowed the time series to periods around the surges by identifying when each variant was known to have first entered North Carolina, based on the retrospective sequencing of clinical samples (29)—from April 5, 2021 onwards for the Delta variant, and from November 29, 2021 onwards for the Omicron variant.

Finally, given our findings from the analyses described above, we developed an algorithm to distinguish signal from noise (underlying variability) and thereby establish an automated way to identify the start of the Delta and Omicron surges. To do so, we considered a combination of several metrics, including the absolute magnitude of flow-population normalized wastewater viral concentrations, sample-to-sample percent changes in those concentrations, and changes in detectability in those concentrations. To assess the performance of our algorithm, we compared algorithmically identified surge start dates with the start dates we had identified based on visual inspection. We then iteratively calculated the performance of different combinations of logical criteria to optimize the algorithm. For these analyses, traditional epidemiologic measures such as sensitivity and the true negative rate are undefined, since all sites had a surge (that is, there was no known negative period, since even prior to entry of the Delta variant, micro-surges in wastewater viral concentrations were observed around holidays and community events).

Instead, we assessed the performance of our algorithm by classifying the number of sites for which the algorithm identified a start date that was (1) within ±1 day of the start date identified through visual inspection (accurate performance), (2) earlier than the visually identified start date (early warning, micro-surge, or false alarm), or (3) later than the visually identified start date (inferior performance). For sites where the algorithm identified an earlier start date, we re-examined the wastewater trend lines in post-hoc fashion to try to explain why an earlier start date was selected. If we found that the algorithm was picking up smaller perturbations in wastewater viral concentrations that were unprecedented but not visually obvious, and which were then closely followed by larger increases, we considered the algorithm to have provided an early warning for the surge (i.e., superior performance). If the algorithm flagged the start of a smaller surge (that is, a series of elevated concentrations that did not reach the levels measured during the main surge), we considered that the algorithm may have flagged a micro-surge (which could have arisen if there were infected individuals shedding viral particles into the sewershed, but whose infections did not lead to widespread community transmission). Lastly, if the algorithm flagged a start date that represented a single or short-term blip that occurred well before precipitous rise and was followed by a period of steady or declining concentrations, we considered that the algorithm likely raised a false alarm (though even in these cases, that single blip could represent a small community outbreak that simply did not lead to widespread or sustained transmission). Based on these criteria, we classified algorithmic performance overall (across both surges) and within each variant-based surge. We also examined how performance changed when we based the algorithm on unnormalized wastewater viral concentrations.

## Data Availability

All data produced are available online at https://covid19.ncdhhs.gov/dashboard/data-behind-dashboards

## Acknowledgements

The authors would like to acknowledge the contributions of several partners to this work, including Shelley Carraway at the Jackson County Department of Public Health; Daniel Manring, Stan Bryson, and Ron Mau at the Tuckaseigee Water & Sewer Authority; Dogwood Health Trust; Rachel Noble, Tom Clerkin, Denene Blackwood, and Rachelle Beattie at UNC-CH; and Melissa Schussman from UWM.

## References

1. A. Keshaviah, X. C. Hu, M. Henry, Developing a Flexible National Wastewater Surveillance System for COVID-19 and Beyond. Environmental Health Perspectives 129, 045002 (2021).

2. C. G. Daughton, Wastewater surveillance for population-wide Covid-19: The present and future. Science of The Total Environment 736, 139631 (2020).

3. K. E. Kapo, M. Paschka, R. Vamshi, M. Sebasky, K. McDonough, Estimation of U.S. sewer residence time distributions for national-scale risk assessment of down-the-drain chemicals. Science of The Total Environment 603–604, 445–452 (2017).

4. S. H. Ali, et al., Regional and socioeconomic predictors of perceived ability to access coronavirus testing in the United States: results from a nationwide online COVID-19 survey. Ann Epidemiol 58, 7–14 (2021).

5. E. N. Asabor, J. L. Warren, T. Cohen, Racial/Ethnic Segregation and Access to COVID-19 Testing: Spatial Distribution of COVID-19 Testing Sites in the Four Largest Highly Segregated Cities in the United States. Am J Public Health 112, 518–526 (2022).

6. H. R. Safford, K. Shapiro, H. N. Bischel, Wastewater analysis can be a powerful public health tool—if it’s done sensibly. Proceedings of the National Academy of Sciences 119, e2119600119 (2022).

7. K. Bibby, A. Bivins, Z. Wu, D. North, Making waves: Plausible lead time for wastewater based epidemiology as an early warning system for COVID-19. Water Res 202, 117438 (2021).

8. E. E. Ooi, J. G. Low, Asymptomatic SARS-CoV-2 infection. The Lancet Infectious Diseases 20, 996–998 (2020).

9. CDC, Instructions for Submitting Human Infection with 2019 Novel Coronavirus (COVID-19) Case Notification Data to CDC Using the CSV Template (2022) (August 20, 2022).

10. M. B. Diamond, et al., Wastewater surveillance of pathogens can inform public health responses. Nature Medicine in press (2022).

11. A. Keshaviah, et al., “The Role of Wastewater Data in Pandemic Management” (2022) (July 26, 2022).

12. P. M. D’Aoust, et al., Quantitative analysis of SARS-CoV-2 RNA from wastewater solids in communities with low COVID-19 incidence and prevalence. Water Res 188, 116560 (2021).

13. J. Peccia, et al., Measurement of SARS-CoV-2 RNA in wastewater tracks community infection dynamics. Nat Biotechnol 38, 1164–1167 (2020).

14. M. Kumar, M. Joshi, A. K. Patel, C. G. Joshi, Unravelling the early warning capability of wastewater surveillance for COVID-19: A temporal study on SARS-CoV-2 RNA detection and need for the escalation. Environ Res 196, 110946 (2021).

15. W. Ahmed, et al., SARS-CoV-2 RNA monitoring in wastewater as a potential early warning system for COVID-19 transmission in the community: A temporal case study. Sci Total Environ 761, 144216 (2021).

16. S. Feng, et al., Evaluation of Sampling, Analysis, and Normalization Methods for SARS-CoV-2 Concentrations in Wastewater to Assess COVID-19 Burdens in Wisconsin Communities. ACS EST Water 1, 1955–1965 (2021).

17. M. Kumar, et al., Lead time of early warning by wastewater surveillance for COVID-19: Geographical variations and impacting factors. Chemical Engineering Journal 441, 135936 (2022).

18. S. W. Olesen, M. Imakaev, C. Duvallet, Making waves: Defining the lead time of wastewater-based epidemiology for COVID-19. Water Research 202, 117433 (2021).

19. A. E. Kirby, Using Wastewater Surveillance Data to Support the COVID-19 Response — United States, 2020–2021. MMWR Morb Mortal Wkly Rep 70 (2021).

20. WRAL, UNC-Charlotte lab confirms state’s first case of coronavirus’ omicron variant: WRAL.com (2021) (August 20, 2022).

21. C. Duvallet, et al., Nationwide Trends in COVID-19 Cases and SARS-CoV-2 RNA Wastewater Concentrations in the United States. ACS EST Water (2022) https://doi.org/10.1021/acsestwater.1c00434 (August 20, 2022).

22. K. E. Graham, et al., SARS-CoV-2 RNA in Wastewater Settled Solids Is Associated with COVID-19 Cases in a Large Urban Sewershed. Environ. Sci. Technol. 55, 488–498 (2021).

23. P. M. D’Aoust, et al., Catching a resurgence: Increase in SARS-CoV-2 viral RNA identified in wastewater 48 h before COVID-19 clinical tests and 96 h before hospitalizations. Sci Total Environ 770, 145319 (2021).

24. R. Noble, et al., “Tracking SARS-CoV-2 in the Wastewater Across a Range of North Carolina Municipalities” (2021) (September 8, 2022).

25. R. E. Beattie, A. D. Blackwood, T. Clerkin, C. Dinga, R. T. Noble, Evaluating the impact of sample storage, handling, and technical ability on the decay and recovery of SARS-CoV-2 in wastewater. PLOS ONE 17, e0270659 (2022).

26. M. Schussman, SARS-CoV-2 RNA Persistence in Municipal Wastewater Treatment Systems Proves Wastewater Surveillance Is an Effective Tool for Monitoring COVID-19 Community Health Burdens. Theses and Dissertations (2022).

27. CDC, National Wastewater Surveillance System. Centers for Disease Control and Prevention (2022) (August 20, 2022).

28. CDC, Wastewater Surveillance Testing Methods (2022) (August 20, 2022).

29. CoVariants, SARS-CoV-2 Mutations and Variants of Interest. SARS-CoV-2 Mutations and Variants of Interest (2022) (August 20, 2022).

