## Supplementary material for "Separating Signal from Noise in Wastewater Data: An Algorithm to Identify Community-Level COVID-19 Surges": Figure S1

### Supplemental Information

#### Figure S1. North Carolina Wastewater Monitoring Network sites analyzed

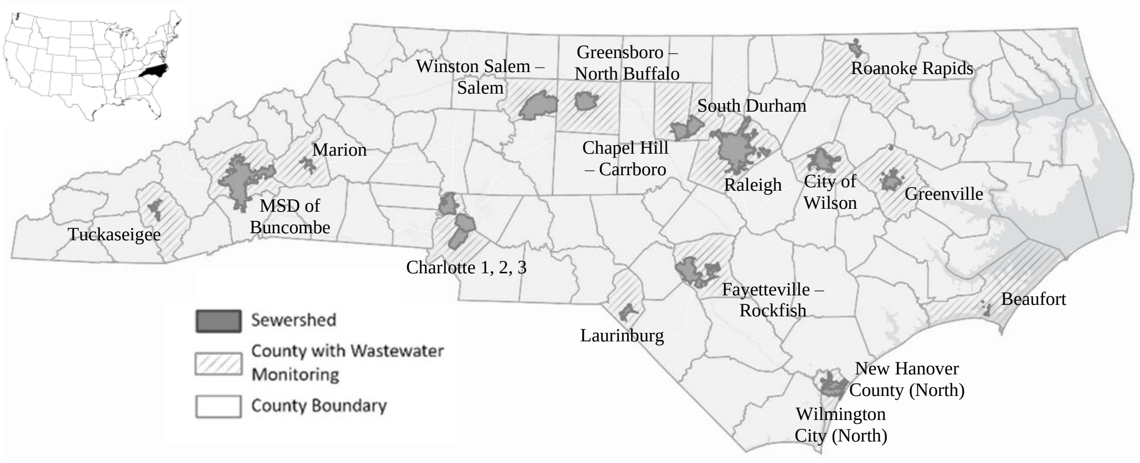


Figure S1. The map shows the sites participating in the North Carolina Wastewater Monitoring Network at the time of the analysis, including the county in which the sites reside and the geospatial boundary of the wastewater treatment plant's service population. Analyses were limited to wastewater treatment plants with sustained monitoring around the Delta and Omicron surges.
