## Supplementary material for "Separating Signal from Noise in Wastewater Data: An Algorithm to Identify Community-Level COVID-19 Surges": Table S1

### Supplemental Information

**Table S1. Correlations between wastewater viral concentrations and unlagged case counts**

| | Spearman rank correlation | | | | |
|---|---|---|---|---|---|
| Site Name | Overall (county-level cases) | Pre-Delta (1/1/2021 to 4/5/2021) | Delta surge (31 days before start of surge to peak of surge) | Omicron surge (31 days before start of surge to peak of surge) | Overall (sewershed-level cases) |
| **Overall (19 sites)** | **0.77** | **0.64** | **0.79** | **0.79** | **0.76** |
| Laurinburg | 0.75* | n/a | 0.76* | 0.72* | 0.74* |
| Tuckaseigee | 0.83* | 0.92* | 0.92* | 0.70* | 0.74* |
| Marion | 0.82* | n/a | 0.65* | 0.88* | 0.81* |
| Beaufort | 0.53* | 0.09 | 0.36* | 0.88* | 0.44* |
| Roanoke Rapids | 0.71* | n/a | 0.82* | 0.72* | 0.72* |
| City of Wilson | 0.80* | n/a | 0.72* | 0.66* | 0.80* |
| Chapel Hill – Carrboro | 0.85* | 0.52* | 0.86* | 0.78* | 0.84* |
| Greenville | 0.86* | 0.73* | 0.85* | 0.79* | 0.85* |
| Wilmington City (North) | 0.81* | 0.63* | 0.88* | 0.83* | 0.77* |
| New Hanover County (North) | 0.56* | 0.61* | 0.75* | 0.55* | 0.53* |
| South Durham | 0.78* | 0.70* | 0.73* | 0.93* | 0.76* |
| Fayetteville – Rockfish | 0.71* | n/a | 0.79* | 0.77* | 0.71* |
| MSD of Buncombe | 0.87* | n/a | 0.94* | 0.88* | 0.86* |
| Winston Salem – Salem | 0.86* | n/a | 0.89* | 0.84* | 0.86* |
| Greensboro – North Buffalo | 0.77* | n/a | 0.81* | 0.80* | 0.74* |
| Charlotte 1 | 0.80* | 0.64* | 0.86* | 0.71 | 0.80* |
| Charlotte 2 | 0.74* | 0.71* | 0.88* | 0.83* | 0.75* |
| Charlotte 3 | 0.79* | n/a | 0.78* | 0.77* | 0.80* |
| Raleigh | 0.88* | 0.86* | 0.74* | 0.92* | 0.86* |

Note: Correlations were calculated between the wastewater viral concentrations (the arithmetic means of the N1 and N2 concentrations, normalized by the flow rate and service population size) and the 7-day rolling average of county-level case counts (unless otherwise specified). Sites are ordered by ascending county population size.
MSD = Metropolitan Sewerage District; n/a = not available (no samples were collected in the timeframe).
*Denotes a correlation coefficient that is statistically significant ($p < 0.05$).
